# Established Machine Learning Matches Tabular Foundation Models in Clinical Predictions

**DOI:** 10.64898/2026.02.02.26345274

**Authors:** Lawrence A. Shaktah, Marco Gustav, Tim Lenz, Junhao Liang, Lars Hilgers, Zunamys I. Carrero, Jakob Nikolas Kather

**Affiliations:** Else Kroener Fresenius Center for Digital Health, Faculty of Medicine, TUD Dresden University of Technology, Dresden, Germany; Department of Medicine I, Faculty of Medicine, TUD Dresden University of Technology, Dresden, Germany; Medical Oncology, National Center for Tumor Diseases (NCT), University Hospital Heidelberg, Heidelberg, Germany; Pathology & Data Analytics, Leeds Institute of Medical Research at St James’s, University of Leeds, Leeds, United Kingdom

**Keywords:** Tabular Data, Machine Learning, Deep Learning, TabPFN, Benchmarking

## Abstract

Foundation models (FMs) promise to standardise predictive modeling across domains, yet their clinical value for tabular data remains unproven. To test this, we performed a large, fully reproducible benchmark of TabPFN, a leading FM for tabular prediction, against twelve established machine learning (ML) methods across twelve binary clinical tasks. Cohorts spanned 788 - 139,528 patients across diverse outcomes, including survival, metastasis, and disease status. Using standardized preprocessing, bootstrapping, and multiple performance metrics, TabPFN was generally competitive but did not consistently outperform strong ML baselines. It exceeded the best ML model in only 16.7% of tasks, with most area under the receiver operating characteristic (AUROC) differences within ±0.01. TabPFN also incurred higher computational cost, with median runtimes 5.5× longer and practical reliance on GPU acceleration. These findings indicate that, for routine clinical tabular prediction, TabPFN offers limited performance gains relative to optimized ML methods, while introducing significant efficiency trade-offs.

## Introduction

Artificial intelligence (AI) and its applications in data science is a rapidly evolving field, and recent advancements have led to the development of large-scale, pre-trained models, referred to as foundation models (FMs). FMs are trained through a self-supervised-learning method^1^, in which the model takes large amounts of unlabeled data and repeatedly tries to learn meaningful patterns that can then be adapted to a wide range of downstream tasks such as classification or prediction^2^. In clinical applications, such models have already been applied to detect meaningful patterns from complex datasets, particularly in domains such as pathology^3^, radiological imaging^4^, and clinical text^5^. The inclusion of FMs in digital pathology workflows has accelerated research^6^ and is beginning to extend into clinical practice. ML models require extensive, and often hand crafted, features to build models from scratch and require task specific labeled data, FM are already embedded with a wide general knowledge base and tend to require less labeled data to fine tune before use; to be as useful if not more useful than any specific ML model. ML models typically require extensive, often hand-crafted, feature engineering and large, task-specific labeled datasets to be constructed from the ground up, whereas FMs are pre-trained on massive, heterogeneous datasets and thus possess a broad, general-purpose knowledge base that permits fine-tuning with far fewer annotations while achieving performance that is comparable to (or surpasses) that of ML models^7^.

Like medical images and clinical text, electronic health record (EHR) data are widely used in healthcare and often capture essential information related to treatments and patient histories. A substantial share of clinical data is tabular, with defined columns such as demographics, laboratory results, and diagnoses that are routinely recorded in this format. An ML or FM trained for this data could therefore offer similar value in clinical settings, as well as new insights, by enabling robust and generalizable predictions across diverse clinical prediction tasks.

A leading foundation model for tabular prediction is Tabular Prior-data Fitted Network (TabPFN), a generative, transformer-based FM introduced and developed by Prior Labs (Prior Labs GmbH, Germany). Hollmann et al. reported that TabPFN outperformed all previous supervised methods for tabular prediction including traditional ML models such as XGBoost, as well as a range of supervised deep learning architectures tailored for tabular prediction such as TabNet, DNF-Net, and FT-Transformer^8^. Unlike ML models that require retraining for each new task, TabPFN performs in-context learning, enabling single-pass predictions without parameter updates. It processes train/test data using a two-way attention mechanism, allowing it to focus on relevant patterns at inference time by capturing relationships across columns. Currently TabPFN is optimized for predictions in small to medium sized datasets, although future versions promise to exceed these limitations^9^. TabPFN was trained on millions of synthetic datasets from varied domains and benchmarked against widely used prediction models, Hollmann et al. reported improvements of ≈ 0.13–0.19 in per-dataset normalized area under the receiver operating characteristic (AUROC) when compared to the strongest baselines^8^. However, TabPFN was trained and evaluated primarily on synthetic tabular data rather than real-world datasets. Given the central role of tabular data in clinical prediction, it remains unclear whether TabPFN can generalize across heterogeneous clinical tasks and cohorts or consistently outperform established ML models. If performance gains are small or inconsistent, the added complexity and computational costs may not justify replacing established tabular ML pipelines.

Previous studies comparing TabPFN with various ML models have reported mixed results. Karabacak et al. conducted twelve independent benchmarking studies on clinical datasets, in which ML models outperformed TabPFN in five cases^12–16^, while TabPFN performed best in three^17–19^, and performance was task dependent in the remaining four^20–23^. At the time of writing, nine additional published studies similarly report heterogenous outcomes, with four favoring TabPFN^24–27^, two ML^28,29^, and three suggesting context-specific performance^30–32^. However, many of these studies rely on private datasets and heterogeneous evaluation pipelines, including differences in feature handling, missing-data strategies, imputation,encoding, class imbalance assessment, resampling, and model tuning. As a result, reported performance differences may reflect pipeline choices as much as underlying model capability.

In this study, we compare established ML models to the TabPFN FM in terms of performance and efficiency to evaluate the prospect of replacing ML models with this new FM. TabPFN was benchmarked against strong representatives of the major model families commonly used for clinical tabular prediction, including classic statistical models^10^, bagging ensembles^11^, boosting methods^12^, and multilayer perceptrons^13^. These families reflect distinct trade-offs in robustness for small-sample tabular data, calibration, interpretability, and computational cost, and together provide a more informative baseline than any single model class. In addition, we quantify runtime on defined hardware and evaluate TabPFN with its recently introduced hyperparameter optimization (HPO) feature. By applying the same pipeline to every target, we obtain performance and efficiency metrics that are directly comparable across models and datasets. This rigorous, uniform framework was essential to isolate genuine differences in model capability from artefacts introduced by heterogeneous analysis pipelines.

## Results

We benchmarked TabPFN against twelve established ML models across twelve binary clinical prediction tasks from eight public datasets, assessing discrimination performance, reproducibility relative to published results, and computational efficiency.

### Benchmark Scope and Task Heterogeneity

The benchmark comprises twelve clinically relevant binary classification tasks derived from three independent data sources: Fundação Oncocentro de São Paulo (FOSP) registry, public datasets curated by Arasteh et al.^40^, and the Surveillance, Epidemiology, and End Results (SEER) cancer registry (Fig. 1A,B). All tasks are based on structured patient-level tabular data with consistent variable representations across datasets (Fig. 1A).

**Figure 1.**
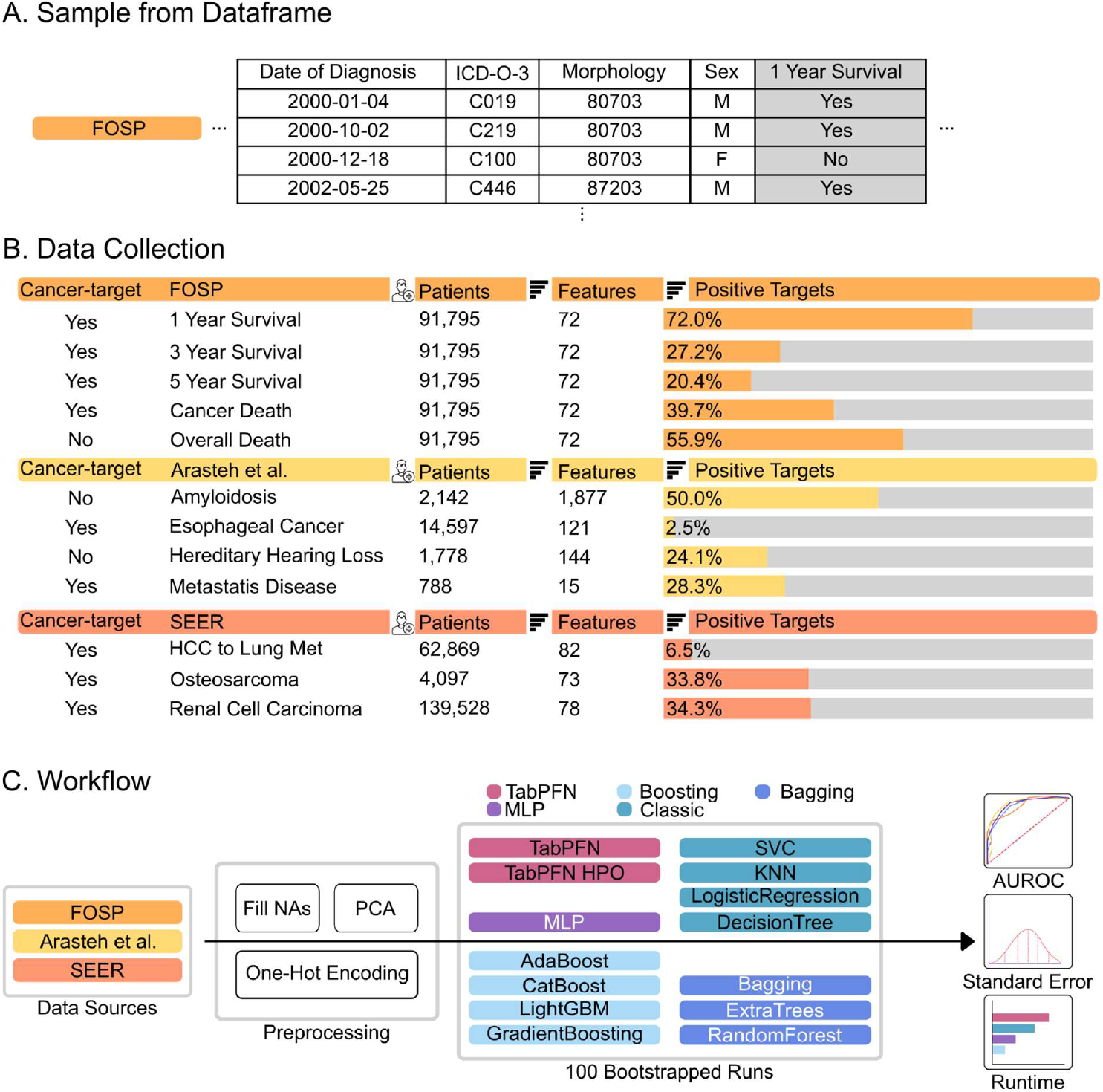
Overview of the TabPFN Benchmark Evaluation Pipeline and Data Sources. (A) Example excerpt from the FOSP dataset, showing patient-level clinical data including diagnosis date, ICD-O-3 code, morphology, sex, and survival outcome. Consistent structured data formats were used across all datasets to establish the binary classification tasks. (B) Patient-level tabular data were collected from three primary sources: the FOSP dataset, the SEER registry, and datasets published by Arasteh et al. These included both cancer-related and non-cancer conditions. Each dataset defines a binary classification task, with target labels representing outcomes such as disease presence or survival at specific time points. (C) All datasets underwent a standardized preprocessing pipeline, including missing value imputation, one-hot encoding, and PCA when feature dimensionality exceeded a prespecified threshold. A comprehensive suite of machine learning models including boosting models, classic algorithms, and TabPFN with and without HPO was trained and evaluated on the processed data. Model performance was assessed over 100 bootstrap iterations using metrics such as AUROC, 95% confidence intervals, and runtime per model iteration.

Across tasks, cohort sizes ranged from 788 to 139,528 patients, feature dimensionality varied from 15 to 1,877 variables, and target prevalences spanned a wide range from rare outcomes (2.5%; esophageal cancer) to common events (72.0%; 1-year survival) (Fig. 1B). The benchmark includes both cancer-related and non-cancer prediction tasks, as well as disease status classification and fixed-horizon survival endpoints.

All datasets were evaluated using a unified benchmarking workflow, including standardized preprocessing, model training across multiple ML families, and repeated bootstrap evaluation with consistent performance metrics (Fig. 1C). This design enables direct comparison of model performance across heterogeneous clinical settings while minimizing confounding effects from dataset-specific evaluation procedures.

### Comparative Discrimination Performance Across Clinical Tasks

Across the twelve clinical prediction tasks, TabPFN achieved discrimination performance comparable to strong ML baselines, with small, task-dependent differences (Fig. 2). When comparing AUROC values for TabPFN, the best-performing ML model, and reported literature benchmarks, all three were closely aligned across tasks (Fig. 2A).

**Figure 2.**
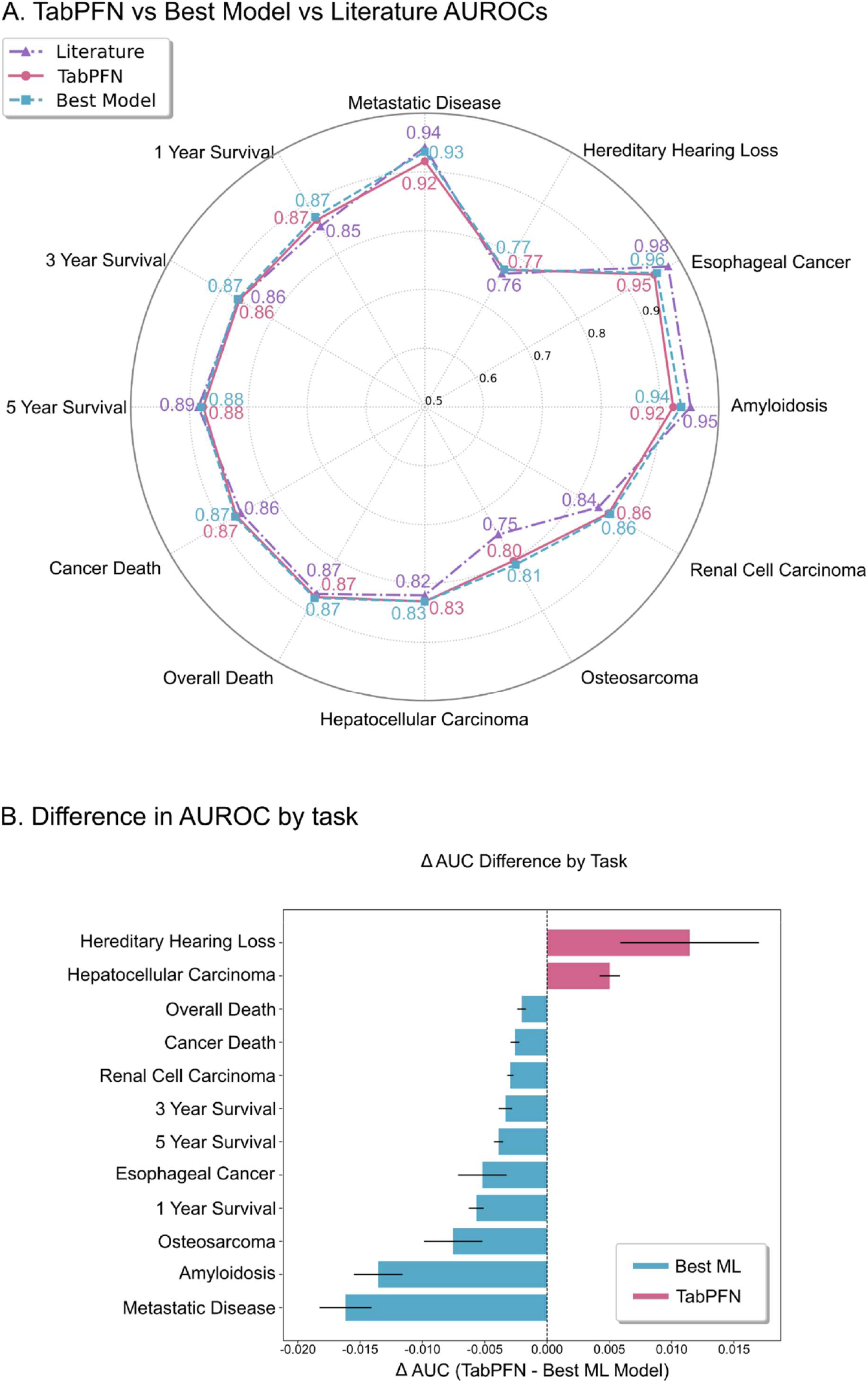
Radar Plot of comparative TabPFN, Machine Learning, and Literature AUROC performance. (A) AUROC across multiple clinical prediction tasks for three benchmarks: TabPFN (pink solid line), the best-performing machine learning model from the benchmark suite (blue dashed line), and reported values from the literature (purple dash-dot line). Prediction tasks span cancer and non-cancer domains, including survival at 1, 3, and 5 years, cancer-specific and overall mortality, as well as disease classification tasks such as Amyloidosis and Hereditary Hearing Loss. TabPFN generally matched the strongest baseline, with small task-dependent differences. (B) Error bars indicate the 95% CI for ΔAUROC derived from the corresponding 95% AUROC CIs. Positive values (pink) indicate superior performance by TabPFN, while negative values (blue) indicate cases where the ML model performs better. In Fig. 2B, ΔAUROC intervals do not cross zero for any task, consistent with the absence of CI-overlap ties.

In direct comparisons against the strongest baseline model per task, TabPFN outperformed the best ML model in 2 of 12 tasks and underperformed in the remaining 10 tasks when considering 95% confidence intervals, with no cases of overlapping intervals. However, the magnitude of these differences was small. Across tasks, most absolute differences in AUROC between TabPFN and the best ML model were within ±0.01 (Fig. 2B), indicating near-parity in discrimination performance.

Both TabPFN and the strongest baseline models, most frequently boosting or bagging approaches such as CatBoost and ExtraTrees, achieved AUROC values close to those reported in the original publications, with only minor deviations observed in isolated cases, including the SEER osteosarcoma task (Fig. 2A). Reported literature AUROCs are included for reference and may not be directly comparable across differing evaluation protocols. Negative-control label permutations yielded AUROC values near 0.5 for all models (Supplementary Figs. S1-S3), confirming the absence of an unfounded signal. Per-model performance summaries are provided in the Supplement (Bagging: S4; Boosting: S5; Classic: S6; TabPFN-HPO and MLP: S7).

### Performance-efficiency Trade-offs

We next examined computational efficiency in relation to discrimination performance (Fig. 3). When AUROC and runtime were normalized to logistic regression as a reference, clear differences emerged across model families in the performance-efficiency plane.

**Figure 3.**
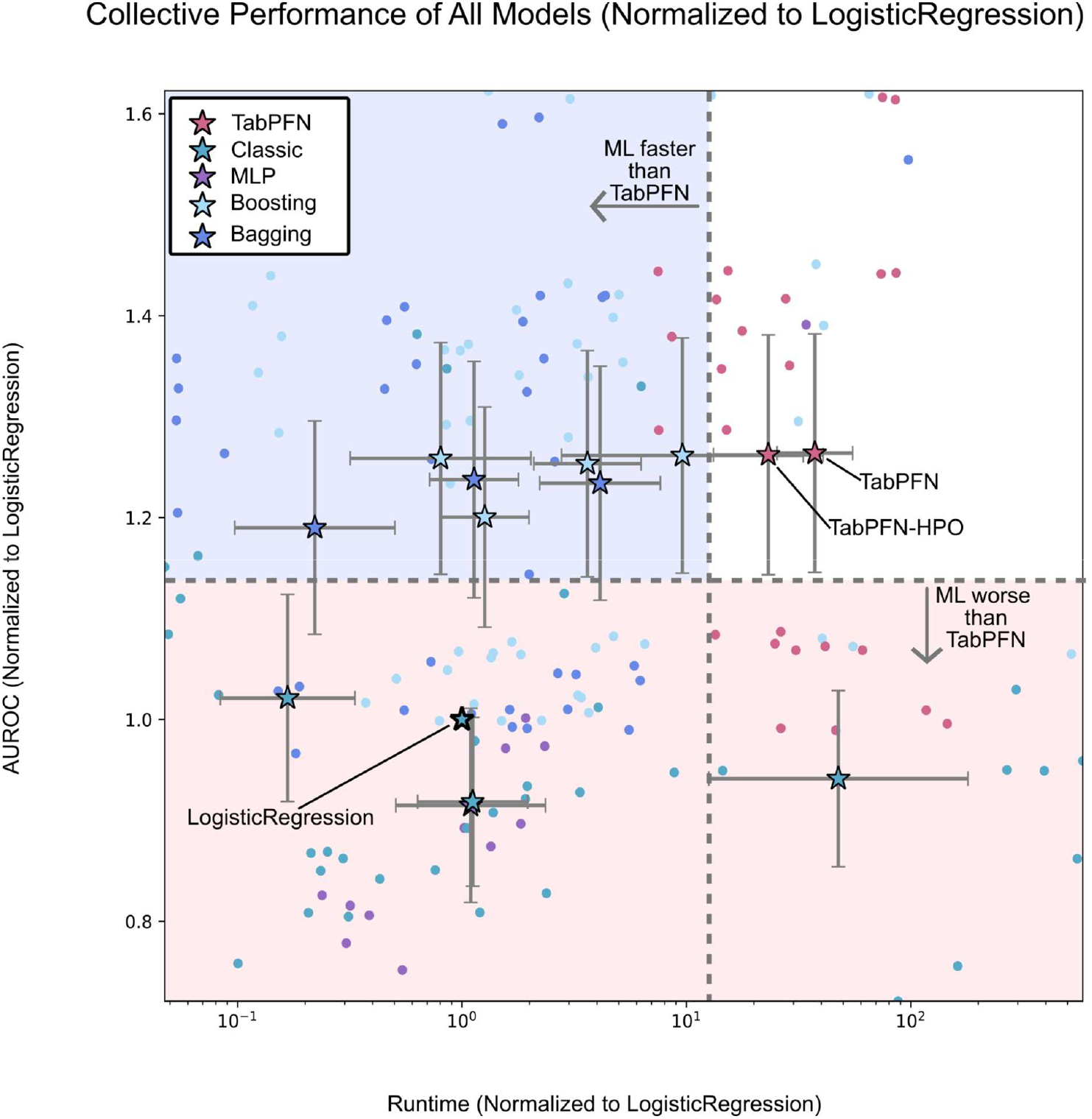
Runtime vs. AUROC normalized to LogisticRegression. Model performance by runtime (x-axis, log scale) and AUROC (y-axis), both normalized to LogisticRegression (reference at [1,1], bolded). Stars represent mean performance for each model class (excluding LogisticRegression reference); TabPFN, boosting, bagging, classic, and multilayer perceptron, all with 95% confidence intervals shown as error bars. Circles denote individual predictions across clinical targets contributing to class means.

Despite achieving AUROC values comparable to the strongest ML baselines, TabPFN exhibited substantially higher computational cost. Across tasks, TabPFN had a median runtime 5.53-fold higher than the best-performing ML model. In contrast, boosting and bagging methods most frequently occupied a favorable region characterized by similar AUROC with markedly lower runtime.

Classic models and multilayer perceptrons generally showed less favorable trade-offs, either due to lower discrimination performance or higher runtime. Across the benchmark, TabPFN did not demonstrate a consistent accuracy advantage sufficient to offset its higher computational cost, placing it in a less favorable region of the performance-efficiency space.

Given TabPFN’s near-parity discrimination with strong ML baselines, we assessed whether comparable performance reflected similar reliance on input features. Permutation SHAP on cardiac amyloidosis prediction revealed a steep long-tailed importance profile for TabPFN, with most attribution mass concentrated in a small subset of predictors (Fig. 4A). Comparing SHAP-based rankings between TabPFN and each baseline showed moderate-to-high agreement, with rank-biased overlap (RBO) values around ∼0.55–0.65 and consistent overlap among the top-20 features (typically ∼7–10 shared features; Fig. 4B). Overall predictive performance was similar, TabPFN generally emphasized the same top predictors as boosting models, with higher computational costs.

**Figure 4.**
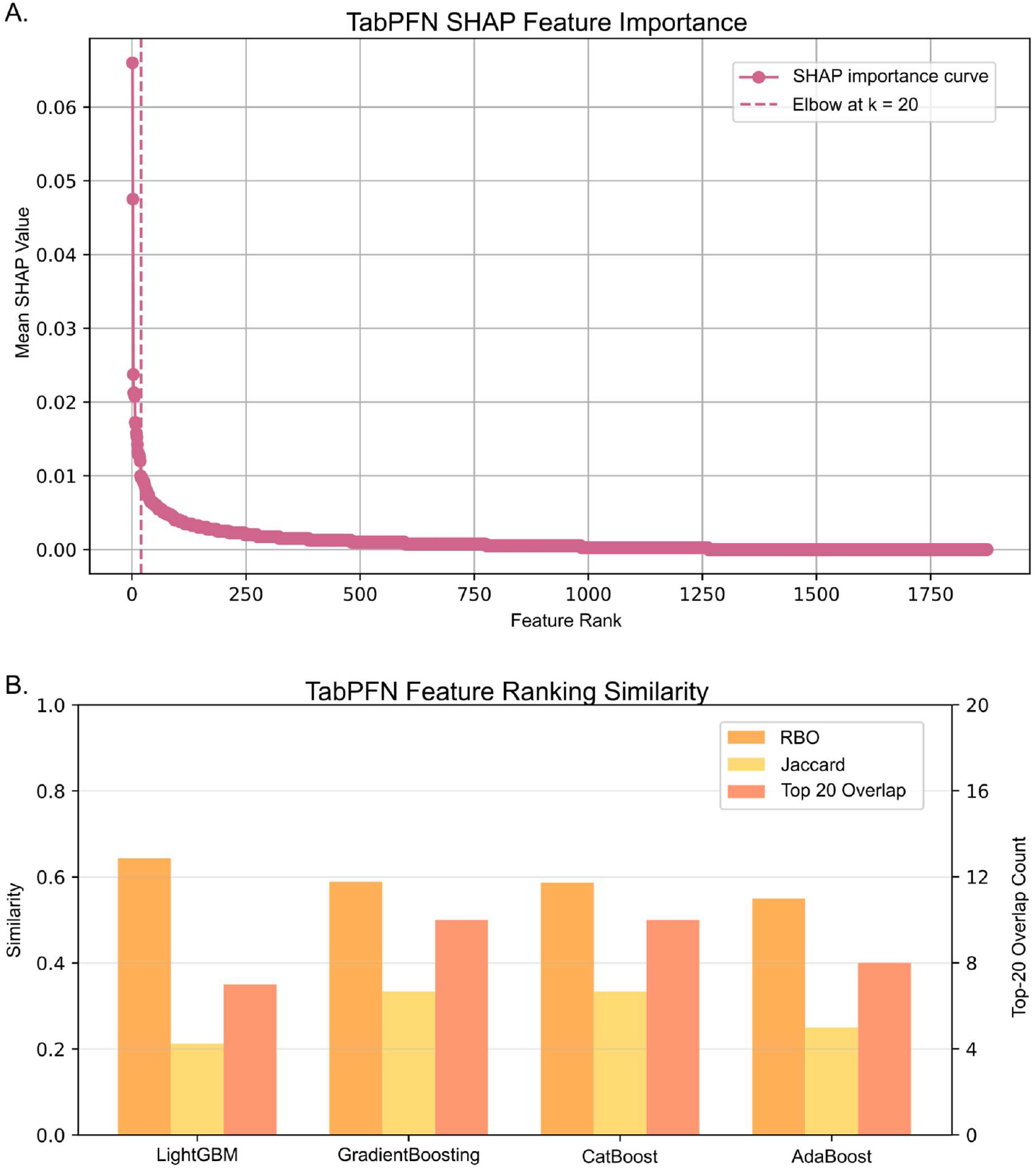
Explainability agreement between TabPFN and classical baselines. (A) Global TabPFN feature-importance curve computed from permutation SHAP. Points show features sorted by decreasing mean absolute SHAP value; the dashed vertical line marks the cutoff k=20 illustrating the rapid drop-off in importance beyond the highest-ranked features. (B) Similarity between TabPFN’s SHAP-based feature ranking and each baseline model’s ranking predicting cardiac amyloidosis. Rank overlap quantified by rank-biased overlap (RBO) (left y-axis indicates similarity among top-ranked features), set overlap by Jaccard similarity on the top-20 features, and absolute agreement by the top-20 overlap count (right y-axis; maximum 20).

## Discussion

We conducted a systematic benchmark of TabPFN against established ML models across a diverse set of clinically relevant tabular prediction tasks, evaluating both its default configuration and its automated HPO. Across tasks, TabPFN achieved discrimination performance comparable to strong ML baselines but did not consistently improve predictive accuracy. In contrast, TabPFN introduced a clear computational trade-off, with substantially longer runtimes than competing approaches. While CPU-only execution was feasible, runtimes were markedly prolonged, whereas bagging and boosting methods frequently achieved similar AUROC values with substantially lower computational cost and without reliance on specialized hardware.

These findings are consistent with prior work showing that ensemble-based methods remain strong baselines for tabular prediction in clinical settings^35^. In addition to competitive performance, boosting and bagging models benefit from mature tooling for feature attribution and error analysis, which facilitates model review and clinical validation^36^. Their relative simplicity also supports deployment on standard CPU-based infrastructure, which remains a practical consideration in many clinical environments.

The absence of a consistent performance advantage for TabPFN may reflect differences between the data regimes used for model development and those encountered in real-world clinical applications. TabPFN was trained primarily on large collections of synthetic datasets, which may not fully capture the heterogeneity, noise structure, and distributional shifts present in clinical tabular data. This interpretation aligns with broader observations in healthcare ML cautioning against replacing well-tuned classical models solely on the basis of architectural novelty^37^.

The observed performance patterns may also reflect practical constraints related to TabPFN’s current row-processing design, which limited the effective use of very large cohorts in this study and may have influenced achievable performance. In addition, the present benchmark focused exclusively on binary classification tasks and did not evaluate multi-class or regression settings, despite TabPFN’s broader capabilities. Future benchmarking efforts that extend to larger tables, additional task formulations, and non-clinical domains will be necessary to more fully assess the potential of tabular foundation models.

Taken together, our results do not support replacing established ML models with TabPFN for routine clinical tabular prediction, particularly when computational efficiency and deployment constraints are critical. In clinical workflows, scalability, interpretability, and operational simplicity are often as important as marginal differences in discrimination performance. Compared with widely used gradient-boosting frameworks such as CatBoost or LightGBM, TabPFN currently offers limited flexibility for domain-specific objectives and constraints. While post-hoc explainability methods can be applied to any model, tree-based ensembles often remain easier to audit in practice due to well-established workflows for attribution and error analysis. In our analysis, TabPFN and Boosting ML models produced similar SHAP-derived importance rankings, implying substantial overlap in the signals each model exploited. Taken together, these results suggest that TabPFN does not consistently offer added benefit for feature discovery or model auditing relative to baselines, at least under post-hoc SHAP attribution analyses.

For foundation models to achieve meaningful adoption in healthcare, they must demonstrate clear and consistent advantages that extend beyond architectural novelty. Although TabPFN may be useful as an out-of-the-box baseline with minimal feature engineering or tuning, broader clinical adoption will depend on improvements in computational efficiency, scalability, and generalizability across diverse clinical tasks and data environments.

## Methods

### Datasets

We constructed twelve clinically relevant binary classification tasks using eight publicly available datasets with established use in the literature. From the Fundação Oncocentro de São Paulo (FOSP), previously used by Woźniacki et al.^38^, we derived five survival-related targets: 1-year survival, 3-year survival, 5-year survival, cancer-related deaths, and overall deaths. FOSP data were acquired from the publicly accessible Fundação Oncocentro Registry^39^, and colorectal cancer cases were identified using ICD-O-3 topography codes C18– C20.

Arasteh et al. provided four tabular clinical prediction datasets for cardiac amyloidosis, esophageal cancer, hereditary hearing loss, and metastatic disease. These datasets were downloaded from the supplementary materials of original benchmark study and used as provided, including predefined splits and engineered features. The tasks corresponded to prediction of amyloidosis risk^41^, hereditary hearing loss inheritance^42^, high-risk esophageal cancer^43^, and cancer metastasis^44^.

From the Surveillance, Epidemiology, and End Results (SEER) program^45^, we reproduced three previously published ML tasks: hepatocellular carcinoma (HCC) metastasis^46^, osteosarcoma (OST)^47^, and renal cell carcinoma (RCC) survival^48^. SEER data were extracted using SEER*Stat (v8.4.5) by replicating published inclusion criteria: (HCC metastasis; “Incidence—SEER Research Data, 17 Registries, Nov 2023 Submission, 2000–2021”; Primary Site C22), (OST survival; “Incidence—SEER Research Data, 17 Registries, Nov 2024 Submission, 2000–2022”; Primary Site C40.0–C41.9), (RCC 5-year overall survival; same 2024 submission; Site recode ICD-O-3/WHO 2008 = Kidney and Renal Pelvis).

All datasets were structured for clinically relevant binary classification tasks.

### Experimental Design

For benchmark comparison, twelve supervised classifiers implemented in *scikit-learn* were evaluated using a fixed random seed for reproducibility. These models included AdaBoost, Bagging, CatBoost, DecisionTree, ExtraTrees, GradientBoosting, K-Nearest Neighbors (KNN), LightGBM, LogisticRegression, RandomForest, Multilayer Perceptron (MLP), and Support Vector Classification (SVC). Hyperparameter settings for all baselineML models are listed in Table 1.

**Table 1.**
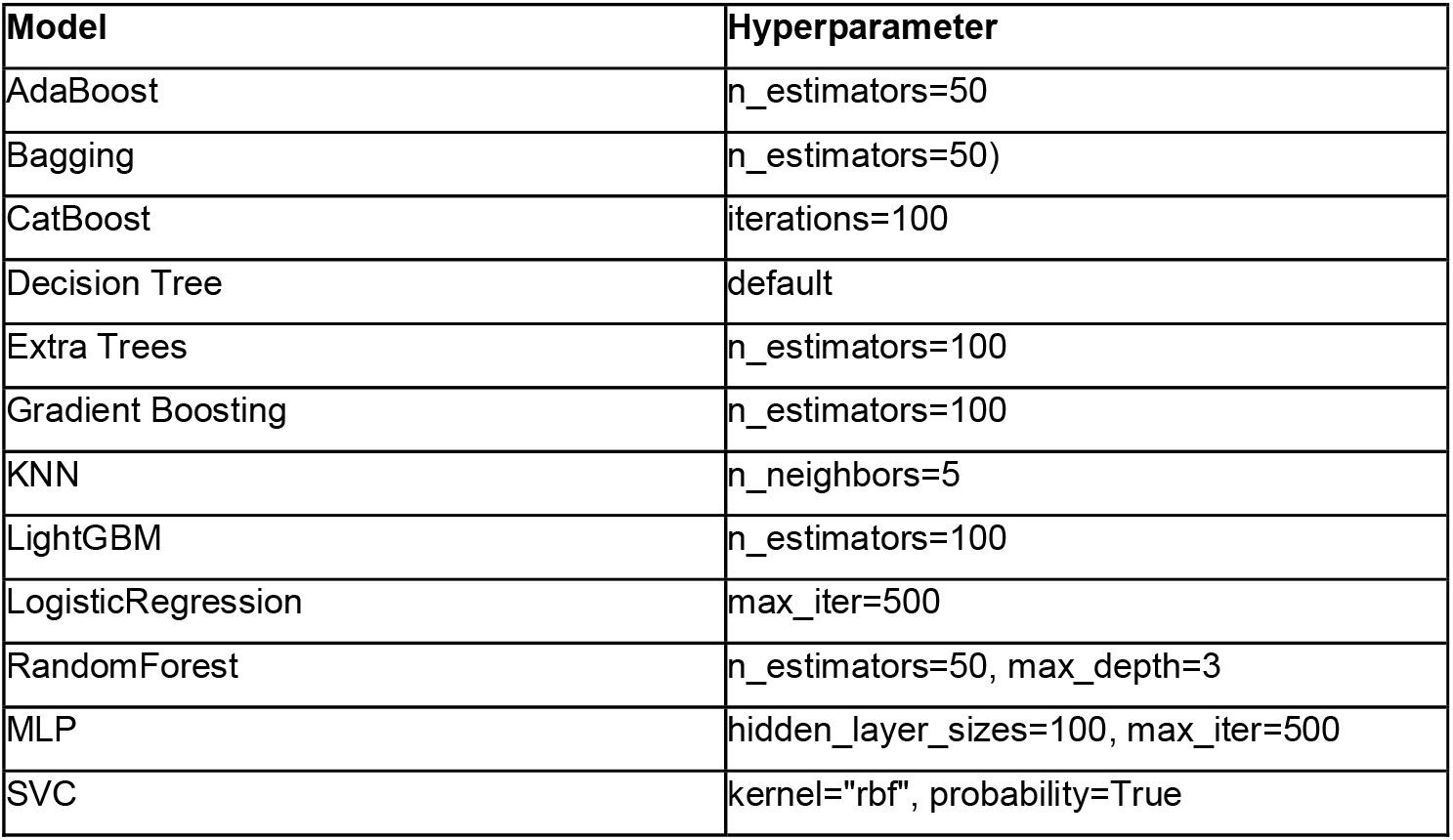
Machine Learning Models and Hyperparameter Settings. This table lists the machine learning algorithms used in the study along with their corresponding hyperparameter configurations. Default parameters were used for models unless specified otherwise.

| Model | Hyperparameter |
| --- | --- |
| AdaBoost | n_estimators=50 |
| Bagging | n_estimators=50) |
| CatBoost | iterations=100 |
| Decision Tree | default |
| Extra Trees | n_estimators=100 |
| Gradient Boosting | n_estimators=100 |
| KNN | n_neighbors=5 |
| LightGBM | n_estimators=100 |
| LogisticRegression | max_iter=500 |
| RandomForest | n_estimators=50, max_depth=3 |
| MLP | hidden_layer_sizes=100, max_iter=500 |
| SVC | kernel="rbf", probability=True |

All experiments were implemented in Python 3.12.7 using TabPFN v2.5 and *tabpfn-extentions* v0.1.2, which supports automated HPO configuration. For each prediction task, TabPFN’s HPO function was allowed 50 iterations to autoselect optimal settings. All models were trained and evaluated on the same features, outcome definitions, and tasks-specific protocols to ensure fair comparison of predictive performance and runtime across models and tasks.

Each clinical target had previously reported AUROC values in the literature, which were reproduced to serve as positive controls. In addition to the default TabPFN configuration, we evaluated the recently introduced TabPFN HPO feature released after the original TabPFN publication^49^.

### General Preprocessing

Across all datasets, minimal preprocessing was applied to maintain comparability. Standard preprocessing steps included missing value handling, categorical encoding, date transformation, outcome engineering, and exclusion of target leakage features.

Missing values in numerical columns were imputed using a constant negative integer and cast to a floating-point format. Missing datetime entries were filled with the placeholder date “1900-01-01”, and missing categorical values labeled as “Missing”. Columns with more than 95% missing values or with (near-)constant (i.e., had ≤1 unique non-null value) were removed. High cardinality categorical columns (>100 unique values) were also excluded to reduce unnecessary model complexity and hinder generalization.

Columns containing date-like strings (YYYY-MM-DD format) were automatically detected and converted to numeric representations as days from 1900-01-01. Target variables and derived outcome-related features, such as “days_survived”, were excluded from all preprocessing steps prior to model training to prevent data leakage. A full list of excluded features is provided in the processing scripts (*Supplemental List 1*).

### Dataset-Specific Preprocessing

FOSP data were filtered to colorectal cancer patients using TOPO codes C18–C20. Outcome variables were defined as follows: overall_death = 1 if ULTINFO = 3 or 4, and cancer_death = 1 if ULTINFO = 3. Survival targets (1-year, 3-year, and 5-year) were computed from DTDIAG and DTULTINFO.

The four datasets from Arasteh et al. (amyloidosis, esophageal cancer, hereditary hearing loss, metastatic disease) were used as provided, including predefined splits and engineered features.

SEER datasets were extracted using disease-specific criteria as described in the Datasets section. HCC data came from the 2023 submission (2000–2021) and were filtered by Primary Site = C22. For OST cases, we used the 2024 submission (2000–2022) with Primary Site =C40.0–C41.9. RCC cases were drawn from the same 2024 submission, filtered using Site recode ICD-O-3/WHO 2008 = Kidney and Renal Pelvis.

### Sampling and Bootstrapping

For each of the twelve clinical prediction tasks, data were preprocessed and features (X) and labels (y) were separated. After one-hot encoding, datasets with more than 2,000 features were reduced via principal-component analysis (PCA) to a 2,000-dimensional subspace, following TabPFN v2.5 input constraints. Data were split into training and testing sets using a 70/30 split.

Model evaluation followed a nested-loop design. In the outer loop, each baseline classifier was evaluated, while the inner loop applied 100 bootstrap resamplings to the training data for each model–target pair. The test set remained fixed. After 100 train/test cycles per model and task, an additional 100 iterations were run as negative controls. We used a fixed stratified split and bootstrapped the training set only; uncertainty therefore reflects sampling variability conditional on the held-out test set.

A second evaluation loop, identical in structure, replaced the baseline models with TabPFN and TabPFN-HPO. Hyperparameters were selected once per task using 50 optimization iterations and then fixed across bootstrap resamplings. All models were evaluated using identical features, targets, and preprocessing. Baseline models were run on CPU, whereas TabPFN models were evaluated using an NVIDIA A100 40GB GPU. AUROC and runtime metrics were collected for all runs.

### Evaluation

Performance metrics included AUROC and runtime, computed per model and per prediction task across 100 bootstrap iterations. Runtimes were measured using system time at the start and end of each loop, then averaged across runs for each model-target pair.

Mean AUROCs with 95% confidence intervals were computed, and TabPFN was compared against both the best-performing baseline and reported literature AUROC on a per-task basis. To assess performance–efficiency trade-offs, AUROC and inference time were normalized to LogisticRegression, enabling direct comparison of model quality versus runtime. No class rebalancing or oversampling was applied, preserving the real-world label imbalance of each dataset.

All threshold-based metrics including balanced accuracy and F1-score, were computed after re-thresholding using the Youden index. Additional metrics, including AUPRC and standard deviations, are reported in Supplemental Table 2.

To compare explainability across models, we computed permutation SHAP for TabPFN and Boosting ML baselines on predicting cardiac amyloidosis using the model’s predicted class as the output. SHAP was estimated on a random sample of up to 20 test instances with a background reference set of up to 1877 features. Global feature importance was summarized as mean absolute SHAP per feature and used to derive ranked feature lists. We quantified agreement between TabPFN and each baseline using RBO, Jaccard similarity on the top 20 features, and the absolute overlap count.

## Data Availability

All datasets analyzed in this study are publicly available.

https://www.nature.com/articles/s41467-021-22876-9

https://github.com/Gaooooye/EsophagealCancer-Screening-Trial

https://data.mendeley.com/datasets/6mh8mpnbgv/1

https://doi.org/10.5281/zenodo.7749613

https://fosp.saude.sp.gov.br/

https://seer.cancer.gov/

## Data Availability

All datasets analyzed in this study are publicly available. The FOSP colorectal cancer registry data were accessed under a data-use agreement with FOSP, using inclusion and exclusion criteria as described in Woźniacki et al. The Amyloidosis, Hereditary Hearing Loss, Esophageal Cancer, and Metastatic Disease datasets were obtained from publicly available sources, with access details provided in the *Methods* section. SEER data were extracted using SEER*Stat version 8.4.5 under a data-use agreement, with derived features described in *Methods*. A full list of datasets and access links is provided in *Supplemental Table 1*.

## Code availability

All code is available at https://github.com/KatherLab/tabpfnbenchmark under an MIT licence for full reproducibility. The code was developed specifically for this study.

## Ethical Consideration

All datasets used in this study were publicly available and fully de-identified prior to access. No new patient data were collected, and no identifiable information was handled at any point. Because all sources are open-access and de-identified, this work did not require institutional ethics approval or informed consent. All data analyses were performed in accordance with the data-use terms of the respective repositories and the principles of the Declaration of Helsinki.

## Author contributions

LAS, MG, LH, and JNK contributed to conceptualization. LAS and JNK contributed to Data Curation. LAS conducted formal analysis. JNK conducted funding acquisition. LAS, MG, LH, JL, and JNK contributed methodology. LAS and JNK contributed to project administration and supervision. LAS, MG, JNK contributed to visualization. LAS, MG, JL, TL, ZIC, and JNK contributed to writing the manuscript.

## Acknowledgments

The authors gratefully acknowledge the Gauss Centre for Supercomputing e.V. (www.gauss-centre.eu) for funding this project by providing computing time through the John von Neumann Institute for Computing (NIC) on the GCS Supercomputer JUWELS at Jülich Supercomputing Centre (JSC). The authors gratefully acknowledge the GWK support for funding this project by providing computing time through the Center for Information Services and HPC (ZIH) at TU Dresden. We would also like to thank Jan Clusmann for helpful discussions and guidance during the initial phase of experimental design. Michaela Unger and Laura Žigutytė for their invaluable support and guidance in our figure designs, as well as Radhika Juglan for her help exploring datasets as we searched for usable targets.

