## Supplementals for "Established Machine Learning Matches Tabular Foundation Models in Clinical Predictions"

#### Supplemental List 1

The following columns were removed from their respective datasets to prevent data leakage.
**Hepatocellular Carcinoma Metastasis:** Combined Summary Stage (2004+), CS mets at dx (2004–2015), CS Mets Eval (2004–2015), Derived EOD 2018 M (2018+), Derived EOD 2018 Stage Group (2018+), EOD Mets (2018+), Histologic Type ICD-O-3, Mets at DX–Distant LN (2016+), Mets at DX–Other (2016+), SEER Combined Mets at DX–bone (2010+), SEER Combined Mets at DX–brain (2010+), SEER Combined Mets at DX–liver (2010+), SEER Combined Mets at DX–lung (2010+), SEER Combined Summary Stage 2000 (2004–2017), SEER historic stage A (1973–2015), Summary stage 2000 (1998–2017), Time from diagnosis to treatment in days recode, Year of diagnosis.
**Osteosarcoma:** COD to site rec KM, COD to site recode, COD to site recode ICD-O-3 2023 Revision, COD to site recode ICD-O-3 2023 Revision, COD to site recode ICD-O-3 2023 Revision Expanded (1999+), SEER cause-specific death classification, SEER other cause of death classification, Survival months, Survival months flag, Vital status recode (study cutoff used).
**Renal Cell Carcinoma:** COD to site rec KM, COD to site recode, COD to site recode ICD-O-3 2023 Revision, COD to site recode ICD-O-3 2023 Revision Expanded (1999+), SEER cause-specific death classification, SEER other cause of death classification, Survival months, Survival months flag, Vital status recode (study cutoff used).

#### Supplemental Figures

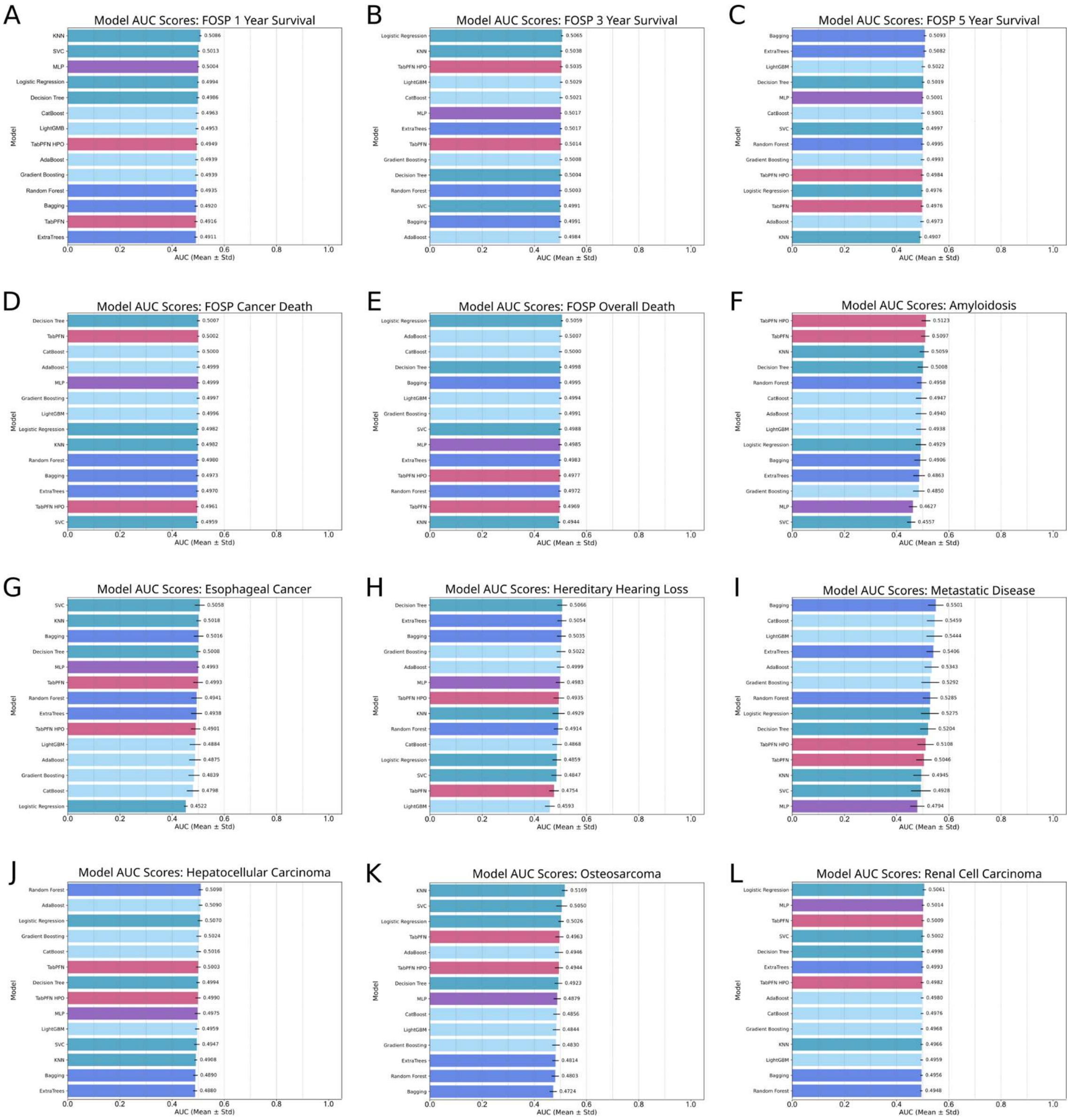


**Supplemental Figure 1. Negative control AUROC performance across FOSP tasks.**

For each target prediction task and corresponding data source, model performance was evaluated using 100 bootstrapped iterations with randomly shuffled outcome labels to serve as negative controls. AUROC scores (mean ± standard deviation) are shown for all models across the following targets: (A) FOSP 1 Year Survival, (B) FOSP 3 Year Survival, (C) FOSP 5 Year Survival, (D) FOSP Cancer Death, (E) FOSP Overall Death, (F) Amyloidosis, (G) Esophageal Cancer, (H) Hereditary Hearing Loss, (I) Metastatic Disease, (J) Hepatocellular Carcinoma, (K) 5 year survival with Osteosarcoma, (L) Renal Cell Carcinoma. All models yielded AUROCs near 0.5, indicating no predictive signal was retained following label permutation, as expected under null conditions.

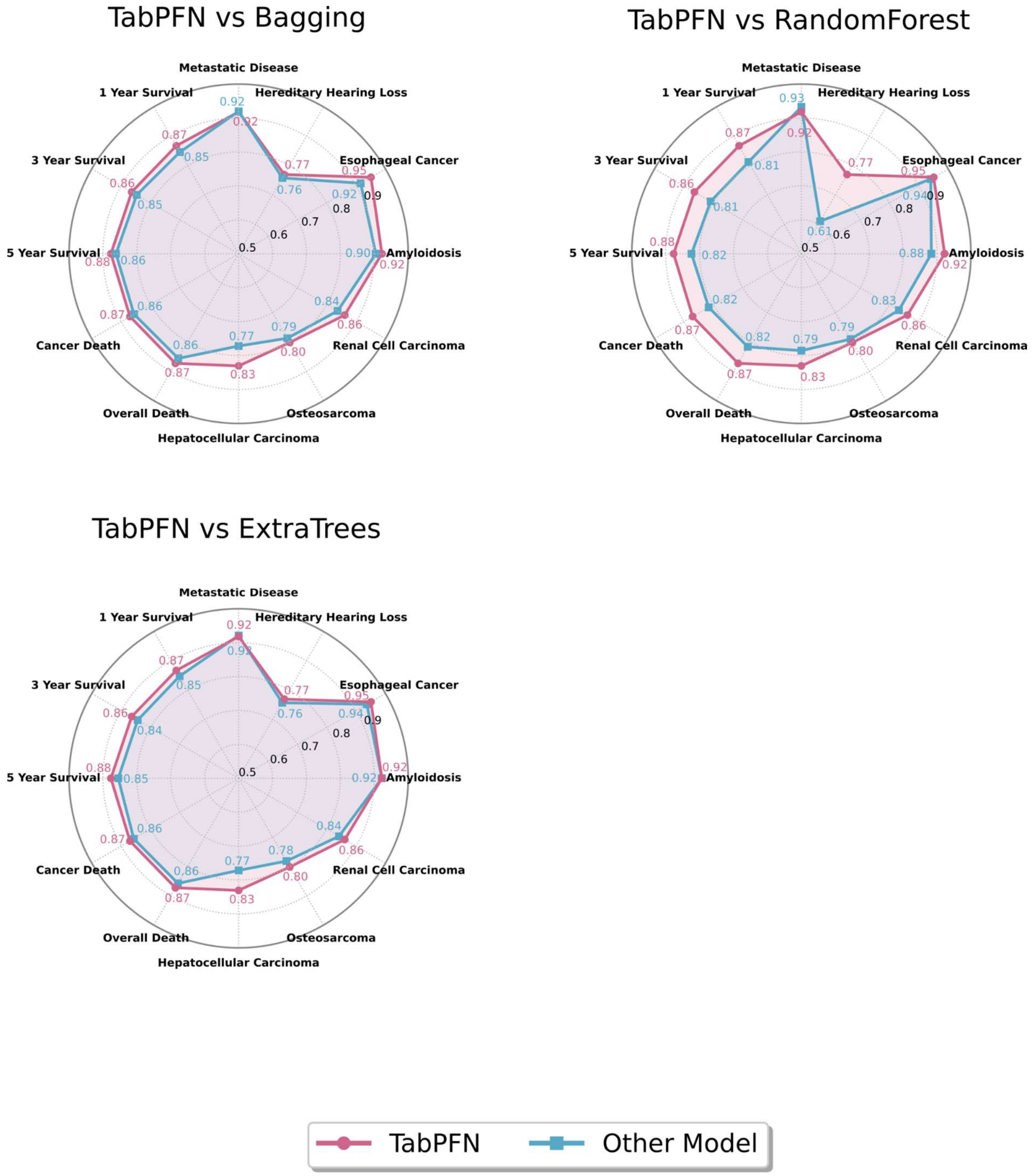


**Supplemental Figure 2. Comparative radar plots of TabPFN versus Bagging ML models across multiple prediction tasks.**

Each radar plot illustrates the performance (measured as AUROC) of TabPFN (blue line) against a different Bagging ML model (red line) across 12 diverse biomedical classification tasks. Each subplot title indicates the comparison. Higher values toward the edge of the radar represent better model performance.

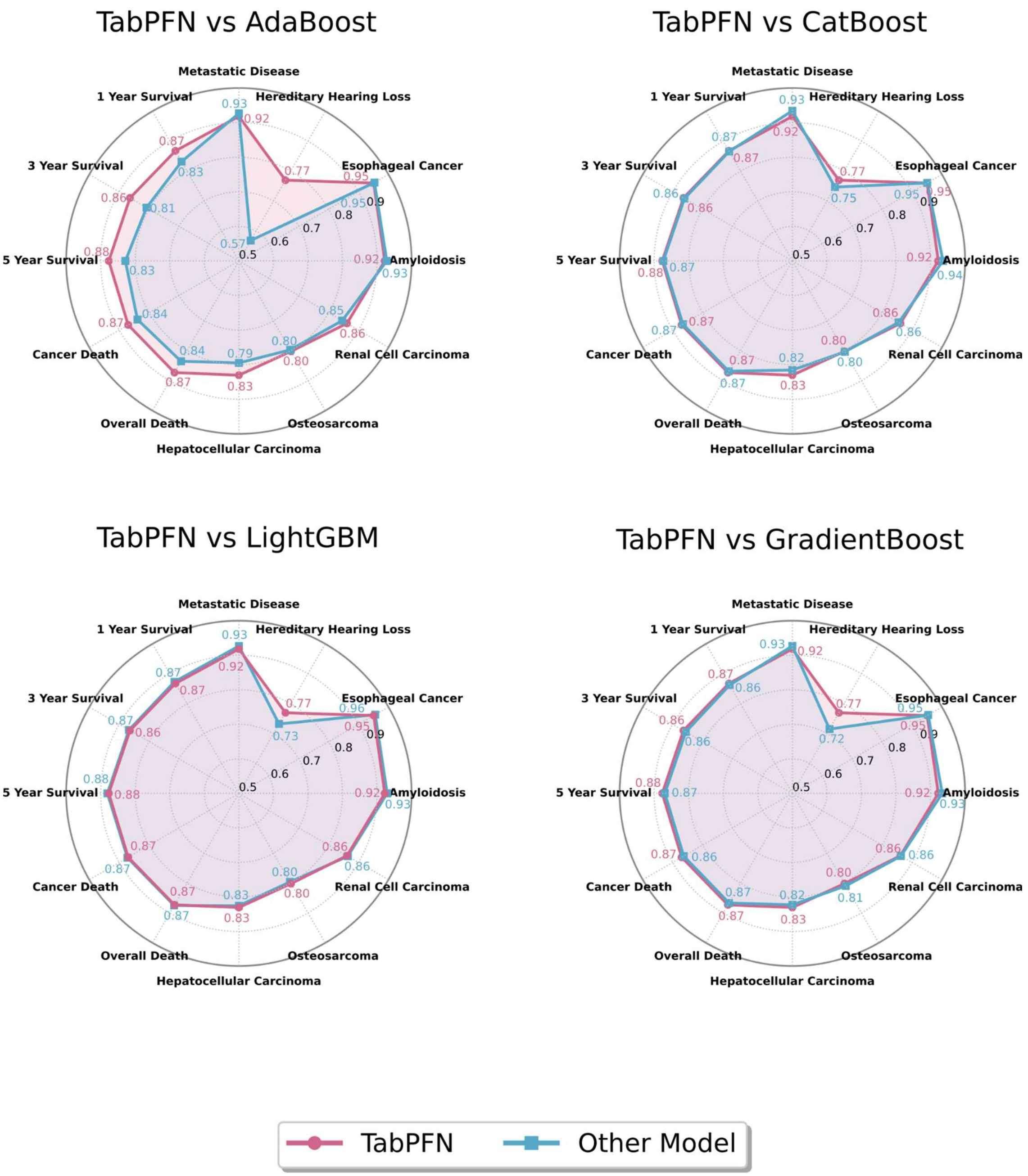


**Supplemental Figure 3. Comparative radar plots of TabPFN versus Boosting ML models across multiple prediction tasks.**

Each radar plot illustrates the performance (measured as AUROC) of TabPFN (blue line) against a different Boosting ML model (red line) across 12 diverse biomedical classification tasks. Each subplot title indicates the comparison. Higher values toward the edge of the radar represent better model performance.

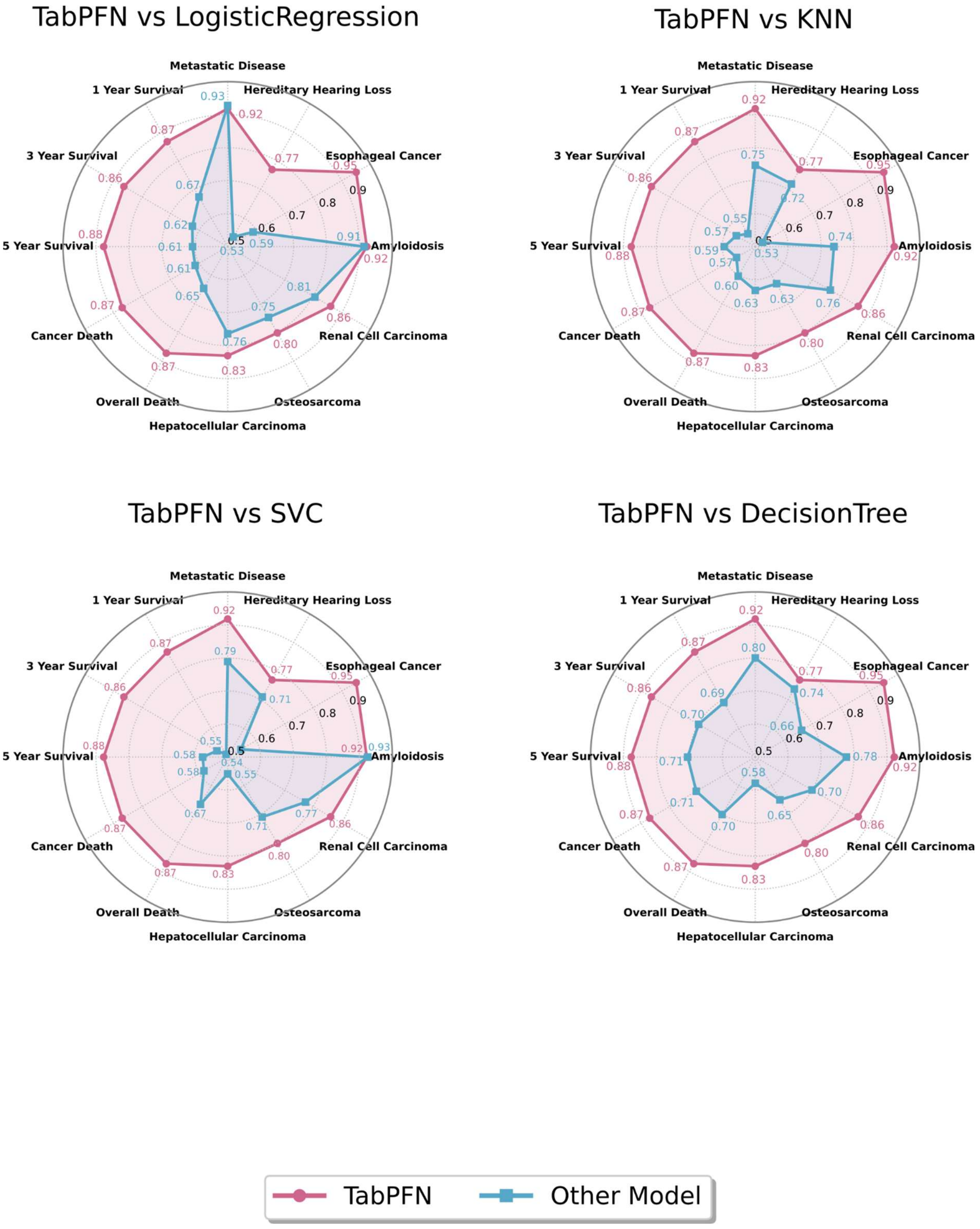


**Supplemental Figure 4. Comparative radar plots of TabPFN versus Classic ML models across multiple prediction tasks.**

Each radar plot illustrates the performance (measured as AUROC) of TabPFN (blue line) against a different Classic ML model (red line) across 12 diverse biomedical classification tasks. Each subplot title indicates the comparison. Higher values toward the edge of the radar represent better model performance.

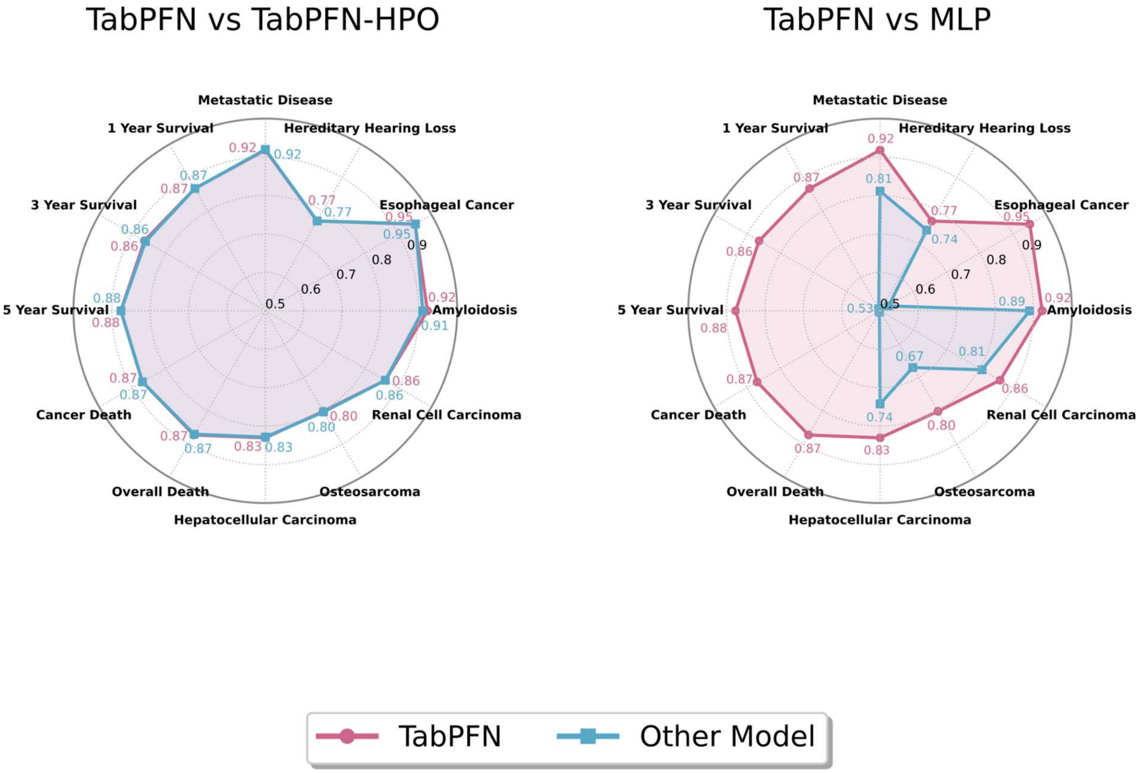


**Supplemental Figure 5. Comparative radar plots of TabPFN versus Neural Net models across multiple prediction tasks.**

Each radar plot illustrates the performance (measured as AUROC) of TabPFN (blue line) against a different Neural Net model (red line) across 12 diverse biomedical classification tasks. Each subplot title indicates the comparison. Higher values toward the edge of the radar represent better model performance.

### Supplemental Table 1

| Target | Source |
|---|---|
| Amyloidosis | https://www.nature.com/articles/s41467-021-22876-9 |
| Esophageal Cancer | https://github.com/Gaoooooye/EsophagealCancer-Screening-Trial |
| Hereditary Hearing Loss | https://data.mendeley.com/datasets/6mh8mpnbgv/1 |
| Metastatic Disease | https://doi.org/10.5281/zenodo.7749613 |
| FOSP (ALL) | https://fosp.saude.sp.gov.br/ |
| SEER (ALL) | https://seer.cancer.gov/ |

**Supplementary Table 1** lists each prediction target and the corresponding public source from which the dataset was obtained (Amyloidosis, Esophageal Cancer, Hereditary Hearing Loss, Metastatic Disease, and the FOSP and SEER registries).

### Supplemental Table 2

| Model | AUROC Mean | AUROFC Std | AUPRC Mean | AUPRC Std | Balanced-Accuracy (Youden) Mean | Balanced-Accuracy (Youden) Std | F1-score(Youden) Mean | F1-score(Youden) Std | Target |
|---|---|---|---|---|---|---|---|---|---|
| LightGBM | 0.8731 | 0.0009 | 0.9515 | 0.0005 | 0.7923 | 0.0012 | 0.8131 | 0.0057 | 1 Year Survival |
| TabPFN | 0.8674 | 0.0022 | 0.9483 | 0.0013 | 0.7899 | 0.0018 | 0.8124 | 0.0051 | 1 Year Survival |
| TabPFN-HPO | 0.8673 | 0.0033 | 0.9484 | 0.0018 | 0.7889 | 0.0029 | 0.8121 | 0.0061 | 1 Year Survival |
| CatBoost | 0.8655 | 0.0015 | 0.9476 | 0.0009 | 0.7861 | 0.0018 | 0.8076 | 0.0061 | 1 Year Survival |
| GradientBoosting | 0.8625 | 0.0007 | 0.9467 | 0.0004 | 0.7825 | 0.0012 | 0.8077 | 0.0047 | 1 Year Survival |
| ExtraTrees | 0.8480 | 0.0011 | 0.9382 | 0.0007 | 0.7716 | 0.0014 | 0.7938 | 0.0064 | 1 Year Survival |
| Bagging | 0.8463 | 0.0016 | 0.9372 | 0.0009 | 0.7688 | 0.0018 | 0.7974 | 0.0068 | 1 Year Survival |
| AdaBoost | 0.8317 | 0.0021 | 0.9341 | 0.0009 | 0.7555 | 0.0021 | 0.7828 | 0.0100 | 1 Year Survival |
| RandomForest | 0.8121 | 0.0032 | 0.9267 | 0.0016 | 0.7329 | 0.0044 | 0.7256 | 0.0138 | 1 Year Survival |
| DecisionTree | 0.6905 | 0.0033 | 0.9100 | 0.0009 | 0.6905 | 0.0033 | 0.7748 | 0.0024 | 1 Year Survival |
| LogisticRegression | 0.6740 | 0.0146 | 0.8610 | 0.0066 | 0.6264 | 0.0112 | 0.6504 | 0.0099 | 1 Year Survival |
| KNN | 0.5452 | 0.0029 | 0.8327 | 0.0012 | 0.5348 | 0.0024 | 0.5636 | 0.0542 | 1 Year Survival |
| SVC | 0.5094 | 0.0171 | 0.7671 | 0.0103 | 0.5171 | 0.0096 | 0.6331 | 0.0870 | 1 Year Survival |
| MLP | 0.5068 | 0.0277 | 0.8743 | 0.0186 | 0.5058 | 0.0219 | 0.1333 | 0.1459 | 1 Year Survival |
| LightGBM | 0.8674 | 0.0007 | 0.8618 | 0.0011 | 0.7852 | 0.0010 | 0.7864 | 0.0010 | 3 Year Survival |
| TabPFN | 0.8641 | 0.0021 | 0.8505 | 0.0266 | 0.7834 | 0.0026 | 0.7845 | 0.0015 | 3 Year Survival |

| | | | | | | | | |
|---|---|---|---|---|---|---|---|---|
| CatBoost | 0.8607 | 0.0012 | 0.8515 | 0.0019 | 0.7811 | 0.0015 | 0.7824 | 0.0015 | 3 Year Survival |
| TabPFN-HPO | 0.8605 | 0.0033 | 0.8472 | 0.0066 | 0.7813 | 0.0021 | 0.7826 | 0.0021 | 3 Year Survival |
| GradientBoosting | 0.8561 | 0.0006 | 0.8467 | 0.0010 | 0.7757 | 0.0009 | 0.7771 | 0.0009 | 3 Year Survival |
| Bagging | 0.8470 | 0.0014 | 0.8363 | 0.0021 | 0.7682 | 0.0018 | 0.7696 | 0.0018 | 3 Year Survival |
| ExtraTrees | 0.8436 | 0.0009 | 0.8317 | 0.0016 | 0.7653 | 0.0014 | 0.7666 | 0.0014 | 3 Year Survival |
| RandomForest | 0.8088 | 0.0030 | 0.7916 | 0.0047 | 0.7399 | 0.0030 | 0.7403 | 0.0031 | 3 Year Survival |
| AdaBoost | 0.8086 | 0.0024 | 0.8067 | 0.0033 | 0.7412 | 0.0028 | 0.7426 | 0.0027 | 3 Year Survival |
| DecisionTree | 0.6986 | 0.0030 | 0.7870 | 0.0021 | 0.6986 | 0.0030 | 0.6991 | 0.0030 | 3 Year Survival |
| LogisticRegression | 0.6239 | 0.0308 | 0.6185 | 0.0291 | 0.5920 | 0.0221 | 0.5927 | 0.0218 | 3 Year Survival |
| KNN | 0.5664 | 0.0026 | 0.6014 | 0.0024 | 0.5485 | 0.0022 | 0.5335 | 0.0090 | 3 Year Survival |
| SVC | 0.5378 | 0.0055 | 0.5319 | 0.0030 | 0.5482 | 0.0053 | 0.5028 | 0.0069 | 3 Year Survival |
| MLP | 0.5029 | 0.0149 | 0.7426 | 0.0420 | 0.5025 | 0.0121 | 0.3192 | 0.0391 | 3 Year Survival |
| LightGBM | 0.8805 | 0.0007 | 0.8008 | 0.0016 | 0.8003 | 0.0012 | 0.7932 | 0.0027 | 5 Year Survival |
| TabPFN | 0.8767 | 0.0012 | 0.7931 | 0.0028 | 0.7973 | 0.0013 | 0.7892 | 0.0026 | 5 Year Survival |
| TabPFN-HPO | 0.8762 | 0.0015 | 0.7914 | 0.0036 | 0.7969 | 0.0016 | 0.7884 | 0.0032 | 5 Year Survival |
| CatBoost | 0.8736 | 0.0011 | 0.7873 | 0.0024 | 0.7951 | 0.0016 | 0.7869 | 0.0033 | 5 Year Survival |
| GradientBoosting | 0.8691 | 0.0006 | 0.7793 | 0.0014 | 0.7884 | 0.0010 | 0.7821 | 0.0031 | 5 Year Survival |
| Bagging | 0.8617 | 0.0013 | 0.7693 | 0.0026 | 0.7843 | 0.0018 | 0.7769 | 0.0034 | 5 Year Survival |
| ExtraTrees | 0.8550 | 0.0010 | 0.7612 | 0.0022 | 0.7772 | 0.0013 | 0.7702 | 0.0038 | 5 Year Survival |

| | | | | | | | | | |
|---|---|---|---|---|---|---|---|---|---|
| AdaBoost | 0.8286 | 0.0015 | 0.7338 | 0.0036 | 0.7562 | 0.0015 | 0.7462 | 0.0071 | 5 Year Survival |
| RandomForest | 0.8239 | 0.0035 | 0.7080 | 0.0068 | 0.7554 | 0.0026 | 0.7481 | 0.0050 | 5 Year Survival |
| DecisionTree | 0.7052 | 0.0029 | 0.7073 | 0.0029 | 0.7052 | 0.0029 | 0.7213 | 0.0026 | 5 Year Survival |
| LogisticRegression | 0.6069 | 0.0232 | 0.4630 | 0.0274 | 0.5827 | 0.0156 | 0.5710 | 0.0245 | 5 Year Survival |
| KNN | 0.5940 | 0.0027 | 0.4713 | 0.0030 | 0.5802 | 0.0020 | 0.4966 | 0.0026 | 5 Year Survival |
| SVC | 0.5760 | 0.0139 | 0.4274 | 0.0069 | 0.5736 | 0.0141 | 0.4846 | 0.0201 | 5 Year Survival |
| MLP | 0.5011 | 0.0077 | 0.6656 | 0.0709 | 0.5011 | 0.0077 | 0.4199 | 0.1042 | 5 Year Survival |
| CatBoost | 0.9360 | 0.0048 | 0.9478 | 0.0033 | 0.8718 | 0.0058 | 0.8712 | 0.0060 | Amyloidosis |
| GradientBoosting | 0.9339 | 0.0052 | 0.9465 | 0.0040 | 0.8706 | 0.0069 | 0.8699 | 0.0071 | Amyloidosis |
| LightGBM | 0.9292 | 0.0052 | 0.9430 | 0.0043 | 0.8674 | 0.0063 | 0.8666 | 0.0065 | Amyloidosis |
| AdaBoost | 0.9281 | 0.0051 | 0.9425 | 0.0037 | 0.8681 | 0.0074 | 0.8674 | 0.0076 | Amyloidosis |
| SVC | 0.9250 | 0.0033 | 0.9394 | 0.0028 | 0.8604 | 0.0058 | 0.8597 | 0.0059 | Amyloidosis |
| TabPFN | 0.9224 | 0.0050 | 0.9336 | 0.0044 | 0.8522 | 0.0072 | 0.8518 | 0.0073 | Amyloidosis |
| ExtraTrees | 0.9224 | 0.0046 | 0.9373 | 0.0040 | 0.8572 | 0.0072 | 0.8566 | 0.0074 | Amyloidosis |
| LogisticRegression | 0.9140 | 0.0052 | 0.9282 | 0.0045 | 0.8465 | 0.0072 | 0.8460 | 0.0072 | Amyloidosis |
| TabPFN-HPO | 0.9102 | 0.0090 | 0.9200 | 0.0097 | 0.8387 | 0.0121 | 0.8383 | 0.0122 | Amyloidosis |
| Bagging | 0.9046 | 0.0083 | 0.9255 | 0.0064 | 0.8506 | 0.0093 | 0.8493 | 0.0097 | Amyloidosis |
| MLP | 0.8899 | 0.0056 | 0.9032 | 0.0052 | 0.8208 | 0.0087 | 0.8203 | 0.0090 | Amyloidosis |
| RandomForest | 0.8833 | 0.0076 | 0.8911 | 0.0087 | 0.8036 | 0.0090 | 0.8033 | 0.0091 | Amyloidosis |

| | | | | | | | | |
|---|---|---|---|---|---|---|---|---|
| DecisionTree | 0.7770 | 0.0173 | 0.8341 | 0.0136 | 0.7770 | 0.0173 | 0.7768 | 0.0173 | Amyloidosis |
| KNN | 0.7391 | 0.0230 | 0.7620 | 0.0226 | 0.6836 | 0.0191 | 0.6795 | 0.0214 | Amyloidosis |
| LightGBM | 0.8726 | 0.0006 | 0.8315 | 0.0010 | 0.7934 | 0.0011 | 0.7989 | 0.0022 | Cancer Death |
| TabPFN | 0.8700 | 0.0012 | 0.8253 | 0.0028 | 0.7920 | 0.0013 | 0.7971 | 0.0025 | Cancer Death |
| TabPFN-HPO | 0.8696 | 0.0023 | 0.8266 | 0.0033 | 0.7909 | 0.0024 | 0.7958 | 0.0031 | Cancer Death |
| CatBoost | 0.8658 | 0.0011 | 0.8196 | 0.0017 | 0.7881 | 0.0014 | 0.7929 | 0.0028 | Cancer Death |
| GradientBoosting | 0.8632 | 0.0005 | 0.8189 | 0.0008 | 0.7859 | 0.0007 | 0.7929 | 0.0018 | Cancer Death |
| ExtraTrees | 0.8569 | 0.0007 | 0.8034 | 0.0014 | 0.7806 | 0.0012 | 0.7856 | 0.0025 | Cancer Death |
| Bagging | 0.8561 | 0.0010 | 0.8087 | 0.0014 | 0.7808 | 0.0013 | 0.7868 | 0.0029 | Cancer Death |
| AdaBoost | 0.8390 | 0.0011 | 0.7822 | 0.0018 | 0.7706 | 0.0019 | 0.7746 | 0.0031 | Cancer Death |
| RandomForest | 0.8155 | 0.0033 | 0.7556 | 0.0036 | 0.7495 | 0.0034 | 0.7548 | 0.0044 | Cancer Death |
| DecisionTree | 0.7068 | 0.0028 | 0.7167 | 0.0027 | 0.7068 | 0.0028 | 0.7188 | 0.0027 | Cancer Death |
| LogisticRegression | 0.6141 | 0.0431 | 0.5043 | 0.0428 | 0.5868 | 0.0314 | 0.5822 | 0.0202 | Cancer Death |
| SVC | 0.5834 | 0.0686 | 0.4637 | 0.0474 | 0.5759 | 0.0276 | 0.5446 | 0.0771 | Cancer Death |
| KNN | 0.5657 | 0.0025 | 0.4704 | 0.0026 | 0.5493 | 0.0025 | 0.5347 | 0.0071 | Cancer Death |
| MLP | 0.5008 | 0.0054 | 0.6766 | 0.0608 | 0.5009 | 0.0055 | 0.3734 | 0.1084 | Cancer Death |
| LightGBM | 0.9559 | 0.0052 | 0.4733 | 0.0269 | 0.9151 | 0.0063 | 0.9341 | 0.0107 | Esophageal Cancer |
| GradientBoosting | 0.9540 | 0.0050 | 0.3564 | 0.0393 | 0.9114 | 0.0053 | 0.9333 | 0.0080 | Esophageal Cancer |
| AdaBoost | 0.9533 | 0.0060 | 0.4101 | 0.0343 | 0.9080 | 0.0074 | 0.9302 | 0.0123 | Esophageal Cancer |

| | | | | | | | | |
|---|---|---|---|---|---|---|---|---|
| TabPFN-HPO | 0.9521 | 0.0042 | 0.5185 | 0.0219 | 0.9154 | 0.0054 | 0.9434 | 0.0075 | Esophageal Cancer |
| CatBoost | 0.9511 | 0.0064 | 0.4841 | 0.0225 | 0.9038 | 0.0078 | 0.9328 | 0.0114 | Esophageal Cancer |
| TabPFN | 0.9507 | 0.0046 | 0.5231 | 0.0220 | 0.9134 | 0.0070 | 0.9406 | 0.0108 | Esophageal Cancer |
| RandomForest | 0.9403 | 0.0039 | 0.3923 | 0.0214 | 0.8969 | 0.0045 | 0.9274 | 0.0124 | Esophageal Cancer |
| ExtraTrees | 0.9365 | 0.0063 | 0.4385 | 0.0203 | 0.8873 | 0.0081 | 0.9271 | 0.0122 | Esophageal Cancer |
| Bagging | 0.9156 | 0.0088 | 0.3793 | 0.0322 | 0.8783 | 0.0085 | 0.9200 | 0.0126 | Esophageal Cancer |
| DecisionTree | 0.6625 | 0.0251 | 0.3107 | 0.0353 | 0.6625 | 0.0251 | 0.9714 | 0.0014 | Esophageal Cancer |
| LogisticRegression | 0.5890 | 0.0086 | 0.0209 | 0.0007 | 0.6443 | 0.0123 | 0.5973 | 0.0408 | Esophageal Cancer |
| SVC | 0.5465 | 0.0502 | 0.0301 | 0.0154 | 0.5627 | 0.0391 | 0.4629 | 0.2295 | Esophageal Cancer |
| MLP | 0.5258 | 0.0329 | 0.1494 | 0.1361 | 0.5261 | 0.0332 | 0.9425 | 0.0896 | Esophageal Cancer |
| KNN | 0.5257 | 0.0108 | 0.0270 | 0.0041 | 0.5262 | 0.0101 | 0.9345 | 0.0106 | Esophageal Cancer |
| TabPFN | 0.8305 | 0.0021 | 0.2709 | 0.0056 | 0.7576 | 0.0026 | 0.7727 | 0.0135 | Hepatocellular Carcinoma |
| TabPFN-HPO | 0.8283 | 0.0022 | 0.2666 | 0.0055 | 0.7554 | 0.0024 | 0.7720 | 0.0166 | Hepatocellular Carcinoma |
| LightGBM | 0.8255 | 0.0021 | 0.2790 | 0.0057 | 0.7497 | 0.0028 | 0.7737 | 0.0167 | Hepatocellular Carcinoma |
| GradientBoosting | 0.8229 | 0.0017 | 0.2685 | 0.0054 | 0.7469 | 0.0025 | 0.7569 | 0.0156 | Hepatocellular Carcinoma |

| | | | | | | | | | |
|---|---|---|---|---|---|---|---|---|---|
| CatBoost | 0.8156 | 0.0032 | 0.2553 | 0.0074 | 0.7434 | 0.0043 | 0.7666 | 0.0177 | Hepatocellular Carcinoma |
| AdaBoost | 0.7950 | 0.0035 | 0.2277 | 0.0062 | 0.7284 | 0.0035 | 0.7355 | 0.0166 | Hepatocellular Carcinoma |
| RandomForest | 0.7855 | 0.0031 | 0.2295 | 0.0044 | 0.7198 | 0.0034 | 0.7128 | 0.0190 | Hepatocellular Carcinoma |
| Bagging | 0.7718 | 0.0042 | 0.2174 | 0.0063 | 0.7149 | 0.0044 | 0.7474 | 0.0224 | Hepatocellular Carcinoma |
| ExtraTrees | 0.7716 | 0.0042 | 0.1923 | 0.0047 | 0.7105 | 0.0043 | 0.7508 | 0.0199 | Hepatocellular Carcinoma |
| LogisticRegression | 0.7641 | 0.0017 | 0.1635 | 0.0023 | 0.7069 | 0.0025 | 0.6967 | 0.0161 | Hepatocellular Carcinoma |
| MLP | 0.7423 | 0.0355 | 0.1567 | 0.0133 | 0.6881 | 0.0261 | 0.7140 | 0.0353 | Hepatocellular Carcinoma |
| KNN | 0.6326 | 0.0048 | 0.1396 | 0.0051 | 0.6298 | 0.0050 | 0.8451 | 0.0017 | Hepatocellular Carcinoma |
| DecisionTree | 0.5794 | 0.0059 | 0.2302 | 0.0098 | 0.5795 | 0.0059 | 0.8919 | 0.0016 | Hepatocellular Carcinoma |
| SVC | 0.5508 | 0.0530 | 0.0836 | 0.0146 | 0.5589 | 0.0315 | 0.6297 | 0.1960 | Hepatocellular Carcinoma |

| | | | | | | | | | |
|---|---|---|---|---|---|---|---|---|---|
| TabPFN-HPO | 0.7701 | 0.0135 | 0.9138 | 0.0063 | 0.7145 | 0.0119 | 0.7546 | 0.0238 | Hereditary Hearing Loss |
| TabPFN | 0.7696 | 0.0124 | 0.9131 | 0.0058 | 0.7158 | 0.0116 | 0.7557 | 0.0222 | Hereditary Hearing Loss |
| Bagging | 0.7581 | 0.0156 | 0.9077 | 0.0079 | 0.7091 | 0.0109 | 0.7479 | 0.0186 | Hereditary Hearing Loss |
| ExtraTrees | 0.7573 | 0.0125 | 0.9071 | 0.0064 | 0.7111 | 0.0101 | 0.7428 | 0.0184 | Hereditary Hearing Loss |
| CatBoost | 0.7466 | 0.0194 | 0.9069 | 0.0090 | 0.7052 | 0.0179 | 0.7376 | 0.0278 | Hereditary Hearing Loss |
| MLP | 0.7428 | 0.0148 | 0.8861 | 0.0125 | 0.7043 | 0.0095 | 0.7559 | 0.0176 | Hereditary Hearing Loss |
| DecisionTree | 0.7377 | 0.0157 | 0.8961 | 0.0075 | 0.7051 | 0.0104 | 0.7532 | 0.0165 | Hereditary Hearing Loss |
| LightGBM | 0.7323 | 0.0134 | 0.9012 | 0.0063 | 0.6979 | 0.0148 | 0.7128 | 0.0271 | Hereditary Hearing Loss |
| KNN | 0.7195 | 0.0271 | 0.8985 | 0.0121 | 0.6827 | 0.0234 | 0.7071 | 0.0611 | Hereditary Hearing Loss |
| GradientBoosting | 0.7151 | 0.0251 | 0.8948 | 0.0118 | 0.6889 | 0.0245 | 0.7100 | 0.0294 | Hereditary Hearing Loss |
| SVC | 0.7102 | 0.0243 | 0.8592 | 0.0171 | 0.7140 | 0.0148 | 0.7617 | 0.0149 | Hereditary Hearing Loss |
| RandomForest | 0.6107 | 0.0347 | 0.8557 | 0.0147 | 0.6205 | 0.0255 | 0.5779 | 0.1216 | Hereditary Hearing Loss |
| AdaBoost | 0.5682 | 0.0178 | 0.8776 | 0.0153 | 0.5758 | 0.0084 | 0.3205 | 0.0884 | Hereditary Hearing Loss |

| | | | | | | | | |
|---|---|---|---|---|---|---|---|---|
| LogisticRegression | 0.5339 | 0.0191 | 0.8113 | 0.0095 | 0.5984 | 0.0114 | 0.4022 | 0.0454 | Hereditary Hearing Loss |
| CatBoost | 0.9343 | 0.0049 | 0.8565 | 0.0140 | 0.8901 | 0.0111 | 0.8943 | 0.0145 | Metastatic Disease |
| RandomForest | 0.9331 | 0.0068 | 0.8573 | 0.0173 | 0.8805 | 0.0101 | 0.8838 | 0.0176 | Metastatic Disease |
| LogisticRegression | 0.9280 | 0.0061 | 0.8467 | 0.0167 | 0.8678 | 0.0071 | 0.8529 | 0.0183 | Metastatic Disease |
| GradientBoosting | 0.9272 | 0.0083 | 0.8559 | 0.0203 | 0.8793 | 0.0116 | 0.8884 | 0.0155 | Metastatic Disease |
| AdaBoost | 0.9269 | 0.0064 | 0.8502 | 0.0148 | 0.8733 | 0.0121 | 0.8761 | 0.0179 | Metastatic Disease |
| LightGBM | 0.9269 | 0.0072 | 0.8578 | 0.0207 | 0.8817 | 0.0106 | 0.8909 | 0.0140 | Metastatic Disease |
| ExtraTrees | 0.9212 | 0.0077 | 0.8370 | 0.0192 | 0.8654 | 0.0102 | 0.8664 | 0.0181 | Metastatic Disease |
| TabPFN-HPO | 0.9200 | 0.0079 | 0.8355 | 0.0163 | 0.8588 | 0.0157 | 0.8654 | 0.0203 | Metastatic Disease |
| Bagging | 0.9199 | 0.0089 | 0.8390 | 0.0233 | 0.8707 | 0.0128 | 0.8787 | 0.0199 | Metastatic Disease |
| TabPFN | 0.9182 | 0.0083 | 0.8338 | 0.0174 | 0.8540 | 0.0149 | 0.8583 | 0.0221 | Metastatic Disease |
| MLP | 0.8113 | 0.0316 | 0.6750 | 0.0359 | 0.7809 | 0.0222 | 0.7855 | 0.0259 | Metastatic Disease |
| DecisionTree | 0.8002 | 0.0313 | 0.7616 | 0.0349 | 0.8002 | 0.0313 | 0.8432 | 0.0223 | Metastatic Disease |
| SVC | 0.7896 | 0.0101 | 0.6244 | 0.0246 | 0.7459 | 0.0118 | 0.7452 | 0.0201 | Metastatic Disease |
| KNN | 0.7468 | 0.0211 | 0.6180 | 0.0308 | 0.6987 | 0.0221 | 0.7308 | 0.0388 | Metastatic Disease |
| GradientBoosting | 0.8093 | 0.0058 | 0.8819 | 0.0060 | 0.7344 | 0.0068 | 0.7410 | 0.0138 | Osteosarcoma |
| TabPFN-HPO | 0.8038 | 0.0059 | 0.8779 | 0.0053 | 0.7360 | 0.0077 | 0.7402 | 0.0142 | Osteosarcoma |
| CatBoost | 0.8036 | 0.0059 | 0.8749 | 0.0056 | 0.7323 | 0.0078 | 0.7368 | 0.0160 | Osteosarcoma |
| TabPFN | 0.8018 | 0.0061 | 0.8772 | 0.0053 | 0.7327 | 0.0075 | 0.7387 | 0.0138 | Osteosarcoma |

| | | | | | | | | | |
|---|---|---|---|---|---|---|---|---|---|
| AdaBoost | 0.7970 | 0.0067 | 0.8721 | 0.0072 | 0.7247 | 0.0070 | 0.7336 | 0.0156 | Osteosarcoma |
| LightGBM | 0.7961 | 0.0063 | 0.8667 | 0.0066 | 0.7293 | 0.0086 | 0.7341 | 0.0161 | Osteosarcoma |
| RandomForest | 0.7904 | 0.0051 | 0.8734 | 0.0045 | 0.7143 | 0.0060 | 0.7304 | 0.0150 | Osteosarcoma |
| Bagging | 0.7874 | 0.0070 | 0.8655 | 0.0059 | 0.7175 | 0.0076 | 0.7308 | 0.0132 | Osteosarcoma |
| ExtraTrees | 0.7819 | 0.0066 | 0.8571 | 0.0062 | 0.7165 | 0.0081 | 0.7245 | 0.0167 | Osteosarcoma |
| LogisticRegression | 0.7476 | 0.0065 | 0.8083 | 0.0079 | 0.6958 | 0.0057 | 0.7226 | 0.0119 | Osteosarcoma |
| SVC | 0.7097 | 0.0188 | 0.8063 | 0.0192 | 0.6673 | 0.0141 | 0.6886 | 0.0261 | Osteosarcoma |
| MLP | 0.6703 | 0.0748 | 0.8125 | 0.0219 | 0.6397 | 0.0587 | 0.6638 | 0.0880 | Osteosarcoma |
| DecisionTree | 0.6497 | 0.0128 | 0.8392 | 0.0058 | 0.6497 | 0.0128 | 0.6806 | 0.0112 | Osteosarcoma |
| KNN | 0.6295 | 0.0133 | 0.7917 | 0.0082 | 0.6009 | 0.0120 | 0.5896 | 0.0376 | Osteosarcoma |
| LightGBM | 0.8749 | 0.0006 | 0.9034 | 0.0006 | 0.7933 | 0.0010 | 0.7908 | 0.0020 | Overall Death |
| TabPFN | 0.8729 | 0.0012 | 0.9010 | 0.0013 | 0.7924 | 0.0014 | 0.7908 | 0.0021 | Overall Death |
| TabPFN-HPO | 0.8707 | 0.0030 | 0.8981 | 0.0034 | 0.7908 | 0.0030 | 0.7889 | 0.0035 | Overall Death |
| CatBoost | 0.8683 | 0.0010 | 0.8968 | 0.0010 | 0.7879 | 0.0013 | 0.7856 | 0.0020 | Overall Death |
| GradientBoosting | 0.8666 | 0.0005 | 0.8956 | 0.0005 | 0.7877 | 0.0008 | 0.7860 | 0.0015 | Overall Death |
| ExtraTrees | 0.8578 | 0.0009 | 0.8873 | 0.0008 | 0.7780 | 0.0013 | 0.7761 | 0.0018 | Overall Death |
| Bagging | 0.8560 | 0.0011 | 0.8852 | 0.0010 | 0.7770 | 0.0017 | 0.7751 | 0.0023 | Overall Death |
| AdaBoost | 0.8350 | 0.0018 | 0.8697 | 0.0012 | 0.7611 | 0.0019 | 0.7586 | 0.0030 | Overall Death |
| RandomForest | 0.8165 | 0.0048 | 0.8524 | 0.0042 | 0.7421 | 0.0056 | 0.7410 | 0.0065 | Overall Death |

| | | | | | | | | | |
|---|---|---|---|---|---|---|---|---|---|
| DecisionTree | 0.7007 | 0.0030 | 0.8099 | 0.0019 | 0.7007 | 0.0030 | 0.7050 | 0.0029 | Overall Death |
| SVC | 0.6654 | 0.0016 | 0.6893 | 0.0017 | 0.6258 | 0.0007 | 0.6337 | 0.0006 | Overall Death |
| LogisticRegression | 0.6462 | 0.0281 | 0.6976 | 0.0234 | 0.6126 | 0.0199 | 0.6032 | 0.0229 | Overall Death |
| KNN | 0.6035 | 0.0026 | 0.6750 | 0.0022 | 0.5773 | 0.0027 | 0.5843 | 0.0026 | Overall Death |
| MLP | 0.5030 | 0.0168 | 0.7722 | 0.0336 | 0.5027 | 0.0139 | 0.3522 | 0.0745 | Overall Death |
| LightGBM | 0.8642 | 0.0006 | 0.9073 | 0.0007 | 0.7869 | 0.0007 | 0.8084 | 0.0026 | Renal Cell Carcinoma |
| GradientBoosting | 0.8631 | 0.0004 | 0.9061 | 0.0006 | 0.7865 | 0.0006 | 0.8076 | 0.0024 | Renal Cell Carcinoma |
| TabPFN | 0.8612 | 0.0007 | 0.9055 | 0.0007 | 0.7841 | 0.0009 | 0.8061 | 0.0028 | Renal Cell Carcinoma |
| TabPFN-HPO | 0.8612 | 0.0013 | 0.9053 | 0.0012 | 0.7843 | 0.0012 | 0.8066 | 0.0027 | Renal Cell Carcinoma |
| CatBoost | 0.8552 | 0.0010 | 0.8979 | 0.0012 | 0.7811 | 0.0011 | 0.8039 | 0.0029 | Renal Cell Carcinoma |
| AdaBoost | 0.8454 | 0.0020 | 0.8892 | 0.0023 | 0.7715 | 0.0018 | 0.7928 | 0.0035 | Renal Cell Carcinoma |
| ExtraTrees | 0.8418 | 0.0009 | 0.8863 | 0.0010 | 0.7710 | 0.0009 | 0.7977 | 0.0030 | Renal Cell Carcinoma |
| Bagging | 0.8370 | 0.0012 | 0.8828 | 0.0012 | 0.7664 | 0.0014 | 0.7914 | 0.0033 | Renal Cell Carcinoma |
| RandomForest | 0.8322 | 0.0013 | 0.8769 | 0.0011 | 0.7688 | 0.0009 | 0.7957 | 0.0021 | Renal Cell Carcinoma |

| | | | | | | | | | |
|---|---|---|---|---|---|---|---|---|---|
| MLP | 0.8070 | 0.0498 | 0.8721 | 0.0137 | 0.7502 | 0.0374 | 0.7784 | 0.0359 | Renal Cell Carcinoma |
| LogisticRegression | 0.8059 | 0.0048 | 0.8432 | 0.0064 | 0.7532 | 0.0014 | 0.7803 | 0.0028 | Renal Cell Carcinoma |
| SVC | 0.7729 | 0.0005 | 0.8347 | 0.0004 | 0.7184 | 0.0005 | 0.7423 | 0.0010 | Renal Cell Carcinoma |
| KNN | 0.7635 | 0.0017 | 0.8608 | 0.0011 | 0.7104 | 0.0020 | 0.7472 | 0.0019 | Renal Cell Carcinoma |
| DecisionTree | 0.6993 | 0.0022 | 0.8603 | 0.0010 | 0.6993 | 0.0022 | 0.7256 | 0.0020 | Renal Cell Carcinoma |

**Supplemental Table 2.** This table presents the performance metrics of top-performing machine learning models for predicting 1 Year Survival. Each metric is reported as the mean and standard deviation across bootstrap samples. The metrics include the Area Under the Receiver Operating Characteristic Curve (AUC), the Area Under the Precision-Recall Curve (AUC-PR), Balanced Accuracy based on the Youden index, and the F1-score at the Youden-optimal threshold. The final column indicates the clinical prediction target corresponding to each model. Grey and white coloring alternate by target class.
